# Policy Implications of an Approximate Linear Infection Model for SARS-CoV-2

**DOI:** 10.1101/2020.06.04.20122549

**Authors:** John E. McCarthy, Bob A. Dumas

## Abstract

We propose a linear model of infection probability, and prove that this is a good approximation to a more refined model in which we assume infections come from a series of independent risks. We argue that the linearity assumption makes interpreting and using the model much easier, without significantly diminishing the reliability of the model.

## 1 Introduction

### 1.1 The need for models for strategic planning

Coronavirus disease 2019 COVID-19, caused by severe acute respiratory syndrome-coronavirus 2 (SARS-CoV-2)], has caused a pandemic. As of June 3, 2020, the World Health Organization reported approximately 6.3 million cases and 380,000 deaths due to the disease WHO. Social distancing and shutting businesses have reduced the number of cases, but there is mounting pressure to reopen businesses. The purpose of this paper is to provide a framework to estimate what the infection risks might be from different activities.

Despite much ongoing research, there are many parameters of coronavirus disease that are not well-understood. Nonetheless policy decisions need to be made despite uncertainty. Some papers have modeled the effects of reopening, e.g. Goldsztejn et al. [2020], Yamana et al. [2020], Chowdhury et al. [2020]. Others have made recommendations on what policies should be implemented, e.g. Miller and Carlson [2020], Pandey et al., Ferretti et al. [2020].

We propose that when planning for activities that last no more than one day, we can use a model of infection probability that is linear in many controllable variables, such as duration of the activity, density of participants, and infectiousness rate among the attendees.

The advantages of a linear model are that it greatly simplifies analyses of different scenarios (for example, the effects of reducing density, or reducing the time spent in specific activities), and also allows comparison of relative risks across different events, even when the base parameters are unknown.

### 1.2 Additivity over time

Suppose we know an upper bound *π* on the chance of an individual becoming infected by SARS-CoV-2 over the course of a day’s activities. Now for some given activity *A*, such as attending a sporting event or taking an airplane flight, we break the activity up into temporally disjoint sub-activities, *S*_1_*, …, S_N_*. (For example: entering the stadium, walking to one’s seat, sitting and watching the event, going to a restroom, leaving the stadium). Suppose the probability of becoming infected in each subactivity *S_j_* is *x_j_*, and we wish to estimate *p*, the probability of becoming infected at some time during *A*. In Appendix 6 we prove that 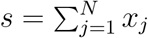 is a good approximation to *p*, and the smaller *π* is, the better the approximation. In particular, we show:

Theorem. *The following inequalities hold:*

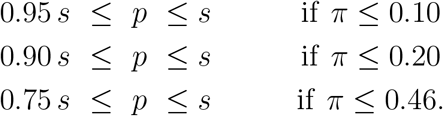

In Subsection 6.2 we give various estimates for *π* with no masks and no social distancing, and they range between 0.18 and 0.47 (this last number corresponds to a doubling time of the number of infected people in the population being just 1.4 days). It seems reasonable to assume that with some awareness of the virus, *π* will be smaller, tightening the estimates in the theorem.

### 1.3 Why is additivity over time important?

If we assume that

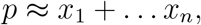

where *≈* means “is approximately equal to”, then it follows that

- A given absolute reduction of risk in any segment *S_j_* has approximately the same overall impact on *p*
- One can compare the relative risks from different activities, such as going grocery shopping, flying, or attending a sporting event, by analyzing sub-activities.

### 1.4 Linearity in other inputs

We describe the full linear model in Section 2.1. In addition to being assumed additive over sub-activities, within each sub-activity it is assumed that the risk of becoming infected is directly proportional to:

- The time spent doing the sub-activity.
- The fraction of attendees who are infectious.
- Density of attendees.
- Transmission probabilities.

In Section 5 we discuss why these are useful when estimating infection probabilities, and hence in planning to keep these low.

## 2 The model

### 2.1 The full linear model

We wish to model the probability that a participant at an activity contracts severe acute respiratory syndrome-coronavirus 2 (SARS-CoV-2). This is understood to happen in one of 3 ways:

- Airborne transmission from an infectious neighbor at the activity.
- Touching a contaminated surface, and then touching the participant’s face before thoroughly washing the hands.
- Direct physical contact with an infectious person.

(A1) Our first assumption is that the model is additive over sub-activities. This means that if one segments the activity into sub-activities (see Subsection 2.3 for an example), the probability of getting infected over the whole activity approximately equals the sum of the probabilities over each segment.

Mathematically, this says that if we break an activity *A* up into *N* distinct sub-activities, *S*_1_*, …, S_N_* say, and the probability *p* of becoming infected during activity *S_j_* is *x_j_* satisfies

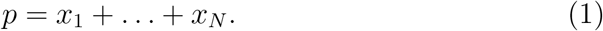

We cannot actually expect exact equality in (1). Nonetheless, and this is an essential point, we can reasonably expect that the left-hand side and right-hand side of (1) agree with each other to within 10% or less. We give a mathematical proof of this assertion in Appendix 6.

(A2) Our second assumption is that for each sub-activity *S_j_* the probability *x_j_* of infection is the sum over the three forms of transmission of three independent probabilities, each of which has a multiplicative form.

For each segment, the probability that the participant becomes infected by air-borne transmission is the sum over every neighbor of [the probability the neighbor is infected] times [the probability the neighbor will cause the participant to be infected per unit time] times [the time spent in their vicinity].

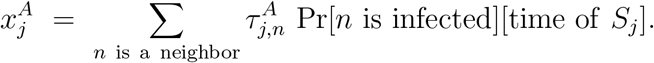

Here 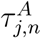 is the probability per unit time that given the configuration (distance away, orientation, mask-wearing or not, etc) that if neighbor *n* is infected, they will infect the participant by air-borne transmission.

The probability that the participant becomes infected by surface-born transmission from a surface they touch is [probability the surface is contaminated] times [probability they touch their face before washing their hands] times [probability that the touching leads to an infection].

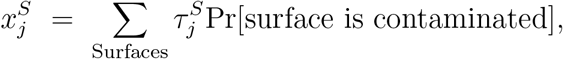

where *τ^S^_j_* is the probability that the participant will convey the infection from the surface to themselves.

The probability that the participant becomes infected by direct contact with an infected neighbor is [probability neighbor is infected] times [probability of touching] times [probability of transmission].

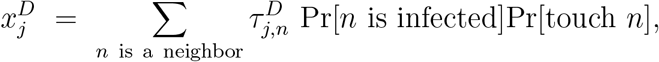

where 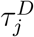 is the probability that if *n* is infected and the participant touches *n*, then infection will be transmitted.

We have

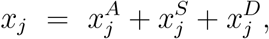

and

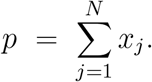

**Our contention is that this model is strategically valuable even without knowledge of the parameters 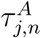, 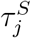, 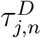.**

### 2.2 Details of the Model

We believe it is very important to have a robust and well-formed, mechanistic, model of infection transmission, even if it contains many unknown parameters. As we will argue in Section 5, this will allow us to draw inferences on *relative risks* even if we cannot quantify *absolute risks*. The availability of a model of infection transmission that is mechanistic, rather than merely statistical, will allow for comparative estimates of the consequences of specific policy choices. Confidence that a policy reduces the risk of infection by a large amount is useful, even in the absence of a reliable estimate of absolute risk.

#### Definitions

By an *activity* we mean a well-defined set of interactions taking place over a period of time less than a day, for example a trip to a grocery store, or taking an airplane flight, or attending a sporting event as a spectator.

By the *participant* we mean a currently non-infected person attending the activity, whose probability of becoming infected we wish to model.

A *neighbor* at an activity is a person not in the participant’s immediate household who, for some part of the activity, is close enough to pose a risk of air-borne infection. We shall say the participant is in the neighbor’s *vicinity* if they are close enough to become infected.

#### Assumptions

We shall make a series of explicit assumptions, in addition to (A1) and (A2) in Subsection 2.1.

(A3) If a neighbor is not infectious, there is 0 risk of infection from them. (A4) If a neighbor is infectious, there is some probability per unit time that they will infect the participant. This probability depends on the distance away, whether they are facing towards or away from the participant, mask usage, viral load in the neighbor, sneeze etiquette, air circulation, and other factors.

(A5) The probability of infection from a neighbor is linear in the amount of time spent in their vicinity. See Appendix 6 for a justification of this assumption.

(A6) The probability of infection in each segment is independent of the other segments.

In this paper we do not need the following assumption, but it will be important when estimating risks of activities.

(A7) There is no increased chance of infection from members of the participant’s immediate household engaging in the same activity, and we will ignore transmission from one’s immediate household members. (So sitting beside a household member at an external activity will be treated as zero-risk).

### 2.3 Segmentation of activities

Each activity is broken down into a sequence of segments, disjoint subactivities each of which can be thought of as a single uniform event, either as a single event (e.g. going to the restroom) or an event with constant parameters (e.g. sitting for some period of time with one neighbor 3 feet away, 2 neighbors 6 feet away, and no other neighbors within 10 feet).

For example, when sitting on a plane, the probability of air-borne infection is the sum over each of the neighboring seats of [the probability that the neighbor is infectious] times [the probability of them infecting the participant per unit time] times [time spent sitting on the plane].

## 3 Examples

To illustrate the utility, we will describe some real-world activities we have analyzed. To do this, we need some numerical form of the decay in risk as people are further apart. So far, this has not been well-established. In the meta-analysis Chu et al. [2020], they found that “A physical distance of more than 1m probably results in a large reduction in virus infection; for every 1m furhter away in distancing, the relative effect might increase 2.02 times”. However, nearly all the papers they studied either compared 0 to 1m, or 1m to 2m, so whether they decay is really exponential is unclear.

### 3.1 Attending a sporting event

We modelled attending a sporting event at a stadium. One can break the event up into three main phases: entering (having one’s ticket checked, going through security, going to one’s seat); sitting for the game; leaving, plus a couple of minor ones (going to the restroom, getting food or drinks).

The utility of the linear model is it allows easy comparison of different scenarios: leaving certain seats vacant; requiring a minimum distance apart in the corridors, so that emptying the stadium takes longer, but there is lower density of foot traffic.

Under all modelled scenarios, the greatest risk by far was from the 3 hours spent watching the game, not entering or exiting, which might be slightly higher risk per minute, but lasted much less time. This suggests that putting plexiglass shields between rows, for example, would make a very bigh difference in the total risk of attending the event.

### 3.2 Taking a flight

This breaks up primarily into

1. Walking through the airport (risk depends on density of people in terminal, and whether mask wearing is required).
2. Going through security
3. Boarding and deboarding plane (risk depends on how this is managed by the airline).
4. Sitting on the plane (risk depends on which seats within 6 feet are occupied by non-household members).

Quantitatively, the risk accrued while sitting on the plane will be much larger than the other risks.

## 4 Limitations of the model

The model depends on many parameters, most of them unknown. We do not know *P r*(infection transmission). This will vary by, among other things,

- Is neighbor wearing a face-mask? What sort?
- Is the neighbor sitting quietly, talking, cheering, singing, moving about?
- What is the neighbor’s cough etiquette?
- Physical proximity and orientation.

We also do not know *P r*(Neighbor is infected). This will vary by, among other things,

- Infection rates in community.
- Willingness of community members to self-isolate if they are symptomatic, or believe they are at risk of being infectious.
- Efficiency of contact tracing and testing to diagnose asymptotatic carriers

We assume in Assumption (A6) that the probability of infection over each segment is independent. This neglects issues of viral load—whether there are threshold levels of exposure at which the probability of infection changes qualitatively.

## 5 Uses of Model

The limitations described in Section 4 mean the model is not useful in calculating absolute risks. Nonetheless, it has significant policy value, as it enables calculations of relative risks. These flow from the fact that the model is linear in several different variables.

1. The model is linear in time. So engaging in an activity for twice as long doubles the chance that a participant becomes infected. Boarding airplanes, for example, can be done in much more efficient ways than is currently the norm Grey. Optimizing the boarding process so that passengers spend less time close to neighbors will reduce their infection risks. Engaging in two successive activities (or two segments of the same activity) the probability of infection is approximately the sum of the probability from each separate activity or segment.
2. The model is linear in the proportion of attendees at an activity who are infectious. This number in turn is the product of two numbers: the proportion of the population who are infectious (which will vary over time) and the probability that an infectious person will not self-isolate. The latter number can be reduced by public health education, by testing and contact tracing, and by health checks.
3. The model is linear in the probability that a given neighbor will cause an infection. In Ma et al. [2020] the authors find that home-made masks block 95% of air-borne viruses, medical masks block 97%, and N95 masks block 99.98%. This was a mechanical simulation using nebulizers and did not distinguish in application between a potentially infectious person wearing a mask to reduce their probability of transmission, and an uninfected person wearing a mask to reduce their probability of infection. In Cheng et al. [2020], the effectiveness of masks in practice is considered. It is reasonable to assume that everybody wearing masks reduces airborne transmission by some factor. In ? they do a meta-analysis, and find that wearing masks reduced risk, with high uncertainty in the amount, but their point estimate was a reduction to 23% of the non-mask wearing risk when using non-respirator masks (and a reduction to 4% using respirators).
4. The model is linear in density of neighbors. Reducing the number of neighbors at a given distance by 50% reduces by 50% the chance of air-borne infection. Leaving seats open, and clustering only members of the same household, will reduce the risk of air-borne transmission.
5. For surface-borne infections, the model is linear in the probability that the surface is contaminated. Doubling the cleaning frequency will approximately halve the probability that the surface is contaminated.

The linearities allow for comparisons among different scenarios, and comparisons across different activities.

## Data Availability

All data used comes from cited sources.

## 6 Appendix: Additivity in time

### 6.1 Mathematical derivation of approximate additivity

Let us assume that an activity *A* is decomposed into *N* segments, called *S*_1_*, …, S_N_*, and each segment *S_j_* has some risk *x_j_* of causing infection. We further assume that these risks are statistically independent of each other. Then the probability *p* of being infected at some time during *A* is

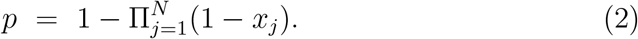

Let

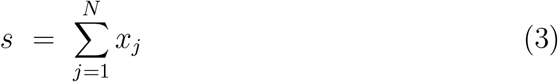

Then we claim that

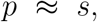

where the symbol *≈* means “is approximately equal to”. Indeed we claim that

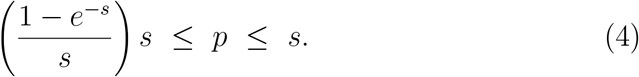

To see (4), we use the arithmetic-geometric inequality to show

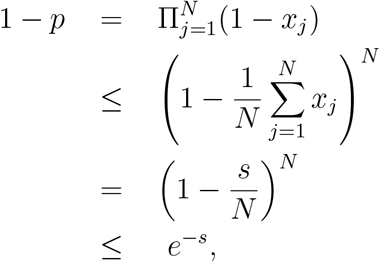

which yields the left-hand inequality in (4).

The right-hand inequality follows from

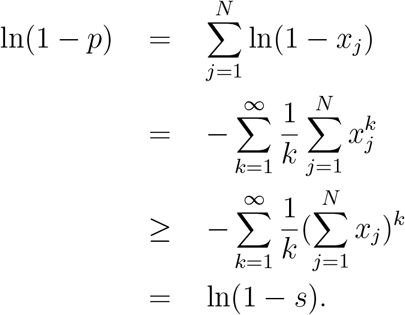

The correction factor between *s* and *p* is 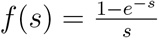. This is a decreasing function of *s*, which has a right-hand limit of 1 as *s* tends to 0. Since *s ≤ −* ln(1 *− p*), we have

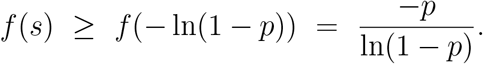

So our conclusion is that we always have

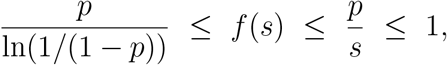

justifying the claim that *p ≈ s*. For representative values of *f*(*s*) and *g*(*p*) = *−p/* ln(1 *− p*), see Tables 1 and 2.

**Table 1:** 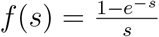 as a function of *s*

|  |  |  |  |  |  |  |  |  |
| --- | --- | --- | --- | --- | --- | --- | --- | --- |
| $s$ | 0 | 0.05 | 0.1 | 0.15 | 0.2 | 0.3 | 0.4 | 0.5 |
| $f(s)$ | 1.00 | 0.98 | 0.95 | 0.93 | 0.91 | 0.86 | 0.82 | 0.79 |

**Table 2:** 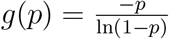 as a function of *p*

|  |  |  |  |  |  |  |  |  |
| --- | --- | --- | --- | --- | --- | --- | --- | --- |
| $p$ | 0 | 0.05 | 0.1 | 0.15 | 0.2 | 0.3 | 0.4 | 0.5 |
| $g(p)$ | 1.00 | 0.97 | 0.95 | 0.92 | 0.90 | 0.84 | 0.78 | 0.72 |

### 6.2 Estimating an upper bound on *p*

As we saw in Subsection 6.1, how good the approximation *p ≈ s* is depends on how close *f*(*s*) is to 1. How can we measure this?

We use the assumption that the activities we are considering last less than a day. While some activities are more risky than others, we further assume that all the events will be designed so that the total risk of an infected individual spreading the infection is no greater than it was at the beginning of the pandemic before any social distancing measures were in place. So we get an upper bound

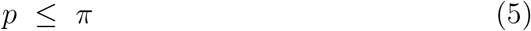

where *π* is the probability that before social distancing, an infectious individual would infect a susceptible individual over the course of one day. Note that for many activities, it is reasonable to assume that *p* is much less than *π*, thus tightening the estimation in Subsection 6.1.

Since an infected individual can infect multiple susceptibles, the expected number they would infect over the course of the day would be slightly higher, namely 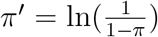if one assumes a Poisson distribution. (This is a small adjustment that will not materially affect our conclusion, so the reader can ignore it.)

Using a standard SIR model at the early stage of an infection, the proportion of the population that is susceptible is close to 1. So the proportion of the population that is infected will grow exponentially, like *e^κt^* for some rate *κ*, where 1 + *π^′^* = *e^κ^*. The doubling time *τ* is the time at which *e^κτ^* = 2, so *κ* = ln(2)*/τ*. Thus we get

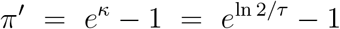

and

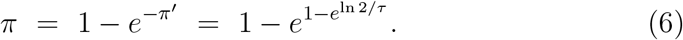

What is *τ*? In Muniz-Rodriguez et al. [2020] they estimate that the doubling times in Chinese provinces in the period January 20 – February 9 2020 ranged from 1.4 days (95% CI 1.2–2.0) in Hunan province to 3.1 days (95% CI 2.1–4.8) in Xinjiang province.

**Table 3:** *π* as a function of *τ*, the doubling time

| $\tau$ | $\pi$ | $f(\pi)$ | $g(\pi)$ |
| --- | --- | --- | --- |
| 1 | 0.63 | 0.74 | 0.63 |
| 1.4 | 0.47 | 0.80 | 0.74 |
| 2 | 0.34 | 0.85 | 0.82 |
| 3 | 0.23 | 0.89 | 0.88 |
| 4 | 0.17 | 0.92 | 0.91 |
| 5 | 0.14 | 0.93 | 0.93 |
| 6 | 0.12 | 0.94 | 0.94 |
| 7 | 0.10 | 0.95 | 0.95 |
| 8 | 0.09 | 0.96 | 0.96 |

In La Maestra et al. [2020], the authors estimate the doubling time in Italy in March 2020 to be 3.4, 5.1 and 9.6 days in the first, second and third ten day periods of the month.

In Karako et al. [2020], the authors estimate the probability of infection in a crowded zone (summed over all neighbors in the vicinity) to be 1.8% per hour, and to be 0.18% and 0.018% in moderate and uncrowded zones, using data from the cruise ship Diamond Princess. If we assume at most 12 hours spent in crowded zones per day on the cruise ship, this would yield the estimate

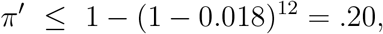

which in turn from (6)gives

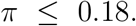

In Berger et al. [2020], the authors use an SEIR model on the data from Kucharski et al. [2020], and take into account the incubation period (6 days) recovery period (14 days) and a mortality rate of 1%. They assume that transmission rates are the same in asymptomatic and symptomatic states, and get a value of *π* that is 0.126. This follows from their equation

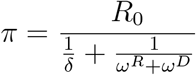

and using their values *R*_0_ = 2.5, *ω^R^* = 1*/*14 is recovery rate from symptomatic to recovered, and *ω^D^* = .01 is the mortality rate.

The value *R*_0_, the number of new people infected per infectious person, is widely reported by time and geographic region – see e.g. Gu for estimates of *R*_0_ by U.S. state. If one makes the more conservative estimate that only asymptomatic carriers will be circulating, and using the same 6 day incubation period, then one gets the bound

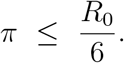

With an estimate of 2.22 for pre-mitigation *R*_0_ in the U.S. Gu, this gives the bound

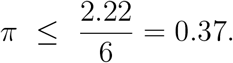

## 7 Appendix: Refinement to additivity over segments

One can refine the analysis in Appendix 6. Let us assume (2) and (3) both hold, and that we have segmented the activity into sufficiently small pieces that each *x_j_ ≤ ε* for some small *ε* that we assume satisfies 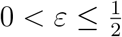.

Then we can tighten the bounds in (4) to

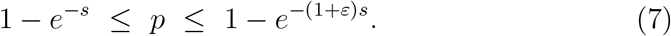

We omit the proof.

## 8 Appendix: Definitions

*π* is the probability that withour social distancing, an infectious individual would infect at least one susceptible individual over the course of one day.

*π^0^* is the expected number of new infections per day caused by an infectious individual without social distancing.

*τ* is the doubling time of the infection.

